# The Generational Health Drift: A Systematic Review of Evidence from the British Birth Cohort Studies

**DOI:** 10.1101/2025.06.12.25329414

**Authors:** Laura Gimeno, Darío Moreno-Agostino, Martin Danka, Yiling Guo, Alice Goisis, Jennifer B Dowd, George B Ploubidis

**Affiliations:** Centre for Longitudinal Studies, Social Research Institute, University College London, London, UK; ESRC Centre for Society and Mental Health, King’s College London, London, UK; Leverhulme Centre for Demographic Science, Nuffield Department of Population Health and Nuffield College, University of Oxford, Oxford, UK

## Abstract

**Background:** Life expectancy improved dramatically during the 20^th^ century. Whether more recent generations are also living longer in good health has serious implications for healthcare systems and the economy.

**Aim:** To synthesise evidence on cohort differences in physical and mental health from the British birth cohort studies, born 1946 to 2000-02.

**Method:** Electronic databases (MEDLINE, EMBASE, PsycInfo, Web of Science, up to 25 June 2024) were searched for pairwise combinations of the six cohort studies of interest or for terms indicating the use of at least two cohorts in the same study. Articles were eligible for inclusion if they compared the health of two or more included cohorts at similar ages (within 3 years).

**Results:** Results were summarised narratively. There was little evidence for improving health across successive cohorts born since 1946 when compared at the same age. For several outcomes – particularly obesity, mental ill-health and diabetes – prevalence of poor health was higher in more recent generations, a pattern we term “Generational Health Drift”. Many outcomes were self-reported, but studies using observer-measured outcomes (anthropometric measures and blood biomarkers) tended to support conclusions based on self-reports.

**Conclusion:** More research is needed to understand the drivers of this trend, shaped by changing exposure to preventable social and environmental risk factors across the lifecourse, and to monitor future trends in disability and functional limitation. The Generational Health Drift has serious implications for policy, planning, and funding allocation to be able to support a growing number of people living with chronic health conditions.

## INTRODUCTION

By 2050, a quarter of the British population will be aged ≥65 years,^1^ with considerable implications for future demand for health and social care and the economy.^2^ Improving health across generations, such that more recently born cohorts live longer in good health, is important to face the challenges of population ageing. Yet reports of stalling increases in life expectancy and health expectancy and persistent health inequalities in the United Kingdom raise questions about whether this improvement is being realised.^3–5^

Whether health is improving across cohorts is not a new question, and has typically been approached using actuarial methods, describing period trends in life and health expectancies.^6,7^ These summary measures of population health are useful for monitoring change over time and quantify the experience of synthetic cohorts constructed using the age-specific mortality rates (and disease prevalence) of a particular year. Period health expectancy in year *x* can be interpreted as the number of years an individual born in year *x* could be expected to live in good health, assuming they experience the mortality and population health conditions of the year of their birth for their entire lives.

In reality, cohorts experience changes in mortality and health conditions during their lifetimes. Cohorts born since the Second World War in Britain have seen remarkable political, social, economic, scientific, and environmental change during their lifetimes.^8^ These changes have affected cohorts at different moments in life and differently shaped their health and mortality risks.^9^ Quantifying changes in health across real-world cohorts is important for policy and planning and complements existing knowledge about trends in life and health expectancies. One approach to this question is to explore whether more recently born cohorts are experiencing a higher prevalence of ill-health compared to previous cohorts at the same age, a trend we refer to as “Generational Health Drift”. This approach offers flexibility since it makes use of prevalence measures, the data for which are routinely collected in population surveys and enables cross-cohort comparisons of health in the absence of mortality data.

The British context is well-suited to exploring trends in health across real-world cohorts, with its collection of long-running birth cohort studies with similar designs, representative of different generations born since 1946.^10–16^ These studies have prospectively collected information on a wide range of health, demographic, and social characteristics across the lifecourse, offering researchers unique opportunities to explore mechanisms underlying changing population health. In this study, we systematically reviewed evidence on age-specific comparisons of physical and mental health across the British birth cohort studies to build a comprehensive picture of how population health is changing across generations.

## METHODS

A protocol was pre-registered with PROSPERO (CRD42024560326; further details in Supplement A).^17^

This review focused on evidence from the 1946 National Survey of Health and Development (1946c),^10^ the 1958 National Child Development Study (1958c),^11^ the 1970 British Cohort Study (1970c),^12,13^ the Avon Longitudinal Study of Parents and Children (ALSPAC; born 1991/92),^14^ Next Steps (born 1989/90),^15^ and the Millennium Cohort Study (2001c; born 2000/02).^16^ These birth cohort studies have followed the lives of people born in different generations (Figure 1, further description in Supplement A).

**Figure 1.**
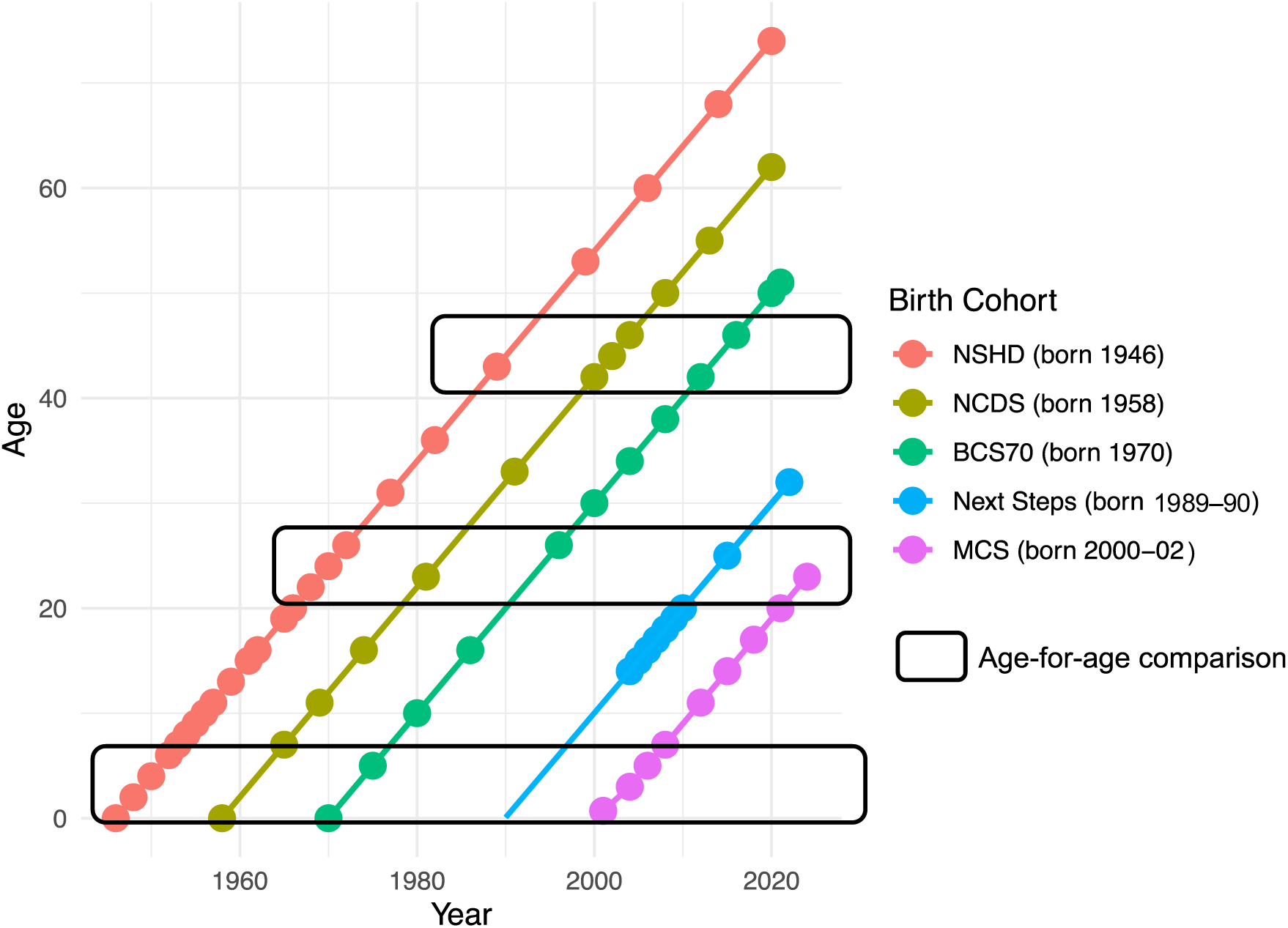
Lexis diagram showing cohort studies included in the review, and data collection sweeps. ***Note:*** *Points represent data collection sweeps. NSHD = 1946 MRC National Survey of Health and Development. NCDS = 1958 National Child Development Study, BCS70 = 1970 British Cohort Study. MCS = Millennium Cohort Study. The Avon Longitudinal Study of Parents and Children (ALSPAC; born 1991/92) is not shown in this graph but closely follows what is shown for Next Steps (born 1989/90), but with more frequent data collection sweeps, starting at birth*.

### Search Strategy and Selection Criteria

Four electronic databases (MEDLINE, EMBASE, Web of Science, PsycInfo, up to 25/06/2024) were searched for pairwise combinations of the six studies, as well for terms indicating the inclusion of two or more British birth cohorts. Search terms for health conditions were not specified to ensure health outcomes were not missed. To be included, studies had to compare health across cohorts when participants were of similar age (within ∼3 years). Inclusion and exclusion criteria are listed in Supplement A. Titles and abstracts and full texts were screened using Rayyan and key bibliographic data and study findings were extracted from included publications.

### Risk of Bias Assessment and Evidence Synthesis

Risk of bias was assessed using the Joanna Briggs Institute Prevalence Studies Criterion,^18^ with attention given to (1) whether analytical samples were representative of their respective birth cohorts at the age of comparison and were derived using comparable selection procedures; (2) how comparable measures of health were in each study; and (3) how the authors addressed attrition and other forms of missing data. Further details can be found in Supplement A, and a list of papers excluded at the full-text screening in Supplement B.

Owing to the variety of outcomes, ages, and combinations of studies compared, we summarised evidence narratively, focusing on the consistency of overall trends for different measures of health across studies, and on the direction of these trends across cohorts (stable, improving, or worsening).

## RESULTS

Database searches yielded 1159 references, of which 51 were deemed eligible (see PRISMA flow diagram in Figure 2). Of these, 37 explicitly compared health or associations between health and other variables across cohorts (Tier 1), and 14 provided descriptive statistics on health outcomes at overlapping ages but did not explicitly aim to compare cohorts (Tier 2).

**Figure 2.**
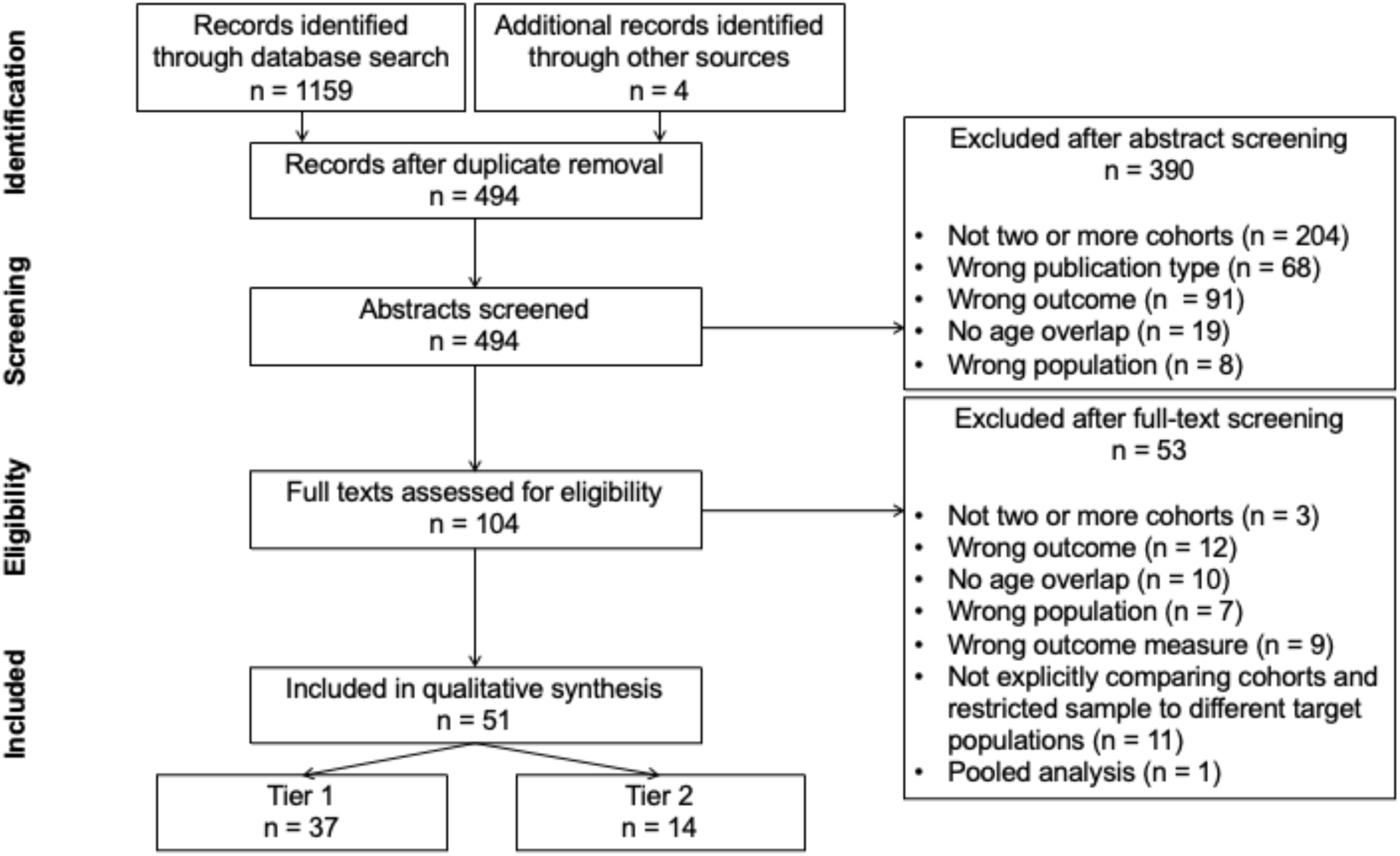
PRISMA Flow Diagram. ***Note:*** *Tier 1 studies compared health or the association between health and other variables across cohorts. Tier 2 studies did not aim to compare cohorts, but present relevant descriptive statistics. Studies could often be excluded for several reasons. We screened papers following the order given in Table S2 and retained the first reason for exclusion as the reason given in* Figure 2.

The most frequently compared outcomes were body mass index (BMI) and obesity (n = 19) and mental ill-health (n = 22). The most frequently compared cohorts were the 1958c and 1970c. Information about included studies and their quality assessment can be found in Supplement C.

### SELF-RATED HEALTH

Self-rated general health (SRH) was compared across the 1958c and 1970c only. Only one study explicitly aimed to compare cohorts, describing mean SRH levels across adulthood, up to age 42 using harmonised measures.^19^ SRH levels were similar across cohorts up to the mid-thirties, when the 1970c experienced a more rapid decline in mean SRH, resulting in a statistically significant difference between cohorts by age 42 (mean score 2.1 versus 2.4, higher score indicating worse SRH). Other studies presenting descriptive statistics generally supported the findings above using different harmonisation approaches, finding small/no differences in poor SRH in cohort members’ thirties,^20–22^ but worse SRH in the 1970c at age 42.^22^

### LIFE SATISFACTION

There was no difference in life satisfaction between the 1958c and 1970c at ages 30/33 or 42.^23–25^ Comparing the 1970c and Next Steps (born 1989-90) at age 25/26, levels of life dissatisfaction were significantly greater in the later-born cohort (20.9% versus 31.5% for men, 19.8% vs. 25.1% for women).^26^

### PHYSICAL HEALTH

#### Anthropometric measures

**Height.** Mean adult height increased across the 1946c, 1958c and 1970c for both men and women,^27,28^ with increases between the 1946c and 1958c appearing to be driven primarily by increasing leg length.^28^ Cohort comparisons of child and adolescent height have additionally included the 2001c. Differences in mean height between the 1946c and 1958c at age 7 were small,^27,28^ but became apparent by age 15/16.^28^ Children in the 1958c grew faster, achieving adult height more quickly.^28^ There was little difference in mean child and adolescent height between the 1958c and 1970c.^29^ The greatest increases in mean height were observed for the 2001c compared to earlier born cohorts (e.g., compared to 1970c, +3.5cm at age 11, +7.5cm at age 15).^29,30^

**BMI, obesity, overweight, and waist and hip circumference.** Mean BMI and overweight/obesity prevalence increased across successive birth cohorts, with cohort differences becoming apparent increasingly early in life.^31^ There was consistent evidence for increases in mean/median BMI and obesity prevalence in midlife (age 42/43) across the 1946c, 1958c and 1970c.^32–39^ Differences in mean childhood/adolescent BMI/obesity prevalence were not apparent between the 1946c and 1958c,^28,29,31,34,40^ with increases in adolescence only becoming apparent with the 1970c, and in childhood for cohorts born after the 1970c,^41^ particularly for the 2001c.^30,42,43^ Cohort differences in BMI/obesity prevalence were reflected in other anthropometric measures, with evidence for increasing mean/median waist and hip circumference in midlife across the 1946c, 1958c and 1970c.^28,32,40^ Studies have also examined cohort differences in the distribution of BMI and waist circumference,^31,32,44^ and in BMI trajectories.^45^

#### Chronic disease

Most evidence came from self-reports of having a particular chronic conditions. For some health conditions (diabetes and high blood pressure), these could be triangulated against biomarker data, yielding insight into cohort changes in both diagnosed and total (diagnosed and undiagnosed) prevalence.

**Diabetes.** The strongest evidence on cohort differences in diabetes prevalence came from comparisons of elevated glycated haemoglobin levels (HbA1c), a blood marker of long-term blood glucose control and a diagnostic criterion for diabetes. Analyses accounting for changes in diabetes medication use found that the prevalence of diabetes in midlife (age 44-48) increased from 3.1% in the 1958c to 5.9% in the 1970c.^37,39^ An increase in prevalence was also observed for self-reported diabetes.^39^ When the 1946c was also considered, there was some suggestion that self-reported diabetes prevalence in cohort members’ thirties had increased across cohorts,^27^ though the number of cases was small and cohort differences were not formally tested. One study quantified self-reported diabetes prevalence in the 1970c and Next Steps (born 1991-92) at age 25/26, but numbers of cases were small and prevalence was reported as <1% in both studies.^43^ Most studies did not differentiate between diabetes type. One study described an increase in diabetes prevalence in childhood (Type 1 diabetes) across the 1946c, 1958c and 1970c,^46^ using information from medical examinations at age 10/11 validated against hospital records. However, the number of cases was small, so prevalence estimates were imprecise.

**High blood pressure.** There was no strong evidence for a consistent cohort trend across the 1946c, 1958c and 1970c using measured systolic and blood pressure in midlife (ages 43/44-45/46-48) even when antihypertensive medication use was accounted for.^33,47^ Prevalence of measured hypertension in midlife was similar in the 1958c and 1970c.^39^ One study comparing the 1946c and 1958c in midlife reported lower mean systolic blood pressure in the 1958c for women only, and lower mean systolic blood pressure for both sexes.^40^ Other studies comparing these two cohorts report different findings, suggesting results may be sensitive to the definition of the analytical cohort is defined and how missing data are addressed.^33,37,47^ Antihypertensive medication use increased dramatically across cohorts.^33,39,40,47^ Only one study examined prevalence of self-reported high blood pressure, reporting similar prevalence in the 1946c and 1958c (for both sexes, age 36/33 and 43/42), a similar prevalence for men and a higher prevalence for women in the 1970c compared to the 1958c (age 30/33). However, cohort differences were not formally tested.^27^

**HDL cholesterol** levels in midlife in the 1858c and 1970c were compared in two studies.^32,37^ Given ongoing debates about the effect of HDL cholesterol on mortality and cardiovascular disease,^48,49^ we refer readers to Supplement A for full discussion of these results.

**Asthma, wheezy bronchitis and bronchitis**. Self-reported asthma/wheezy bronchitis has been compared in the 1946c, 1958c, 1970c, ALSPAC and 2001c. Prevalence was generally higher in more recently born cohorts. Lifetime prevalence of asthma/wheezy bronchitis in midlife was higher in the 1970c than the 1958c,^39^ with differences in both lifetime prevalence and period prevalence already apparent at age 16.^50^ 12-month asthma prevalence was significantly higher in 2001c compared to the 1970c at age 10/11.^30^ The prevalence of wheeziness in the last year in ALSPAC and 2001c was similar across multiple reports between birth and age 10,^51^ though it is worth noting that there were differences in attrition rates between studies, and members of ALSPAC are more socioeconomically advantaged than those of the 2001c. Comparisons including the 1946c were more challenging because question wording differed. One study found that asthma prevalence increased across the 1946c, 1958c and 1970c (at ages 36/33/30 and 43/42), while trends for bronchitis prevalence were more mixed, decreasing between the 1946c and 1958c at age 42/43, but with no clear trend across the 1946c, 1958c and 1970c at age 36/33/30 (cohort differences were not formally tested).^27^

**Hay fever and eczema**. Two studies described cohort differences in hay fever and three in eczema prevalence. The 12-month prevalence of hay fever increased significantly between the 1958c and 1970c at age 16,^52^ and between the 1970c and 2001c at age 10/11.^30^ Eczema prevalence also increased across cohorts, with 12-month prevalence at age 16 increasing from 3.2% in the 1958c to 6.2% in the 1970c,^52^ and was higher in the 2001c than the 1970c at age 10/11.^30^ Lifetime prevalence of eczema in the 1970c by age 42 was 28%, compared to 18% in the 1958c by age 50.^53^

**Cancer**. Three studies compared lifetime prevalence of cancer. There was no difference between 1958c and 1970c at age 46-48 (1% in both cohorts).^39^ Comparisons at younger ages were challenging given the small number of individuals with the outcome. Age-specific comparisons of lifetime cancer prevalence in the 1946c and 1958c at age 43/42, the 1958c and 1970c at age 33/30,^27^ and the 1970c and Next Steps at age 25/26 (among men),^43^ all reported similar prevalences across cohorts. Comparisons were not disaggregated by type, which could obscure diverging trends for specific cancer types.

**Other chronic conditions**. Three studies provided the only evidence on cohort differences in longstanding illness (1970c/1958c at 30/33),^20^ back pain (1946c/1958c/1970c at 36/33/30; 1946c/1958c at 43/42),^27^ and migraines, epilepsy, and multimorbidity (1958c/1970c at 42).^39^ Prevalence of all outcomes was higher in later-born cohorts. An exception was back pain, for which there was no clear trend and question wording varied markedly between studies.^27^

#### Limiting longstanding illness, sensory impairment, and cerebral palsy

Three studies compared the prevalence of longstanding limiting illness the 1958c and 1970c (at 33/30 and 42), finding a slightly higher prevalence in the 1970c with greater differences for women (though differences were not formally tested).^20,22,27^ One study compared the 1970c and 2001c (at 10/11), finding no difference in childhood limiting longstanding illness prevalence.^30^ Cross-cohort comparisons of visual and hearing impairment were limited.

Prevalence of glasses use at age 10/11 was slightly higher in the 2001c than the 1970c, while prevalence of parent-reported hearing problems decreased slightly.^30^ Although long- and short-distance unaided vision tests using similar protocols were conducted in the 1946c, 1958c and 1970c at ages 10/11 and 15/16, the two studies using this data were limited by large amounts of missing data.^54,55^ One study on cerebral palsy in the 1958c and 1970c found that prevalence at birth was similar across cohorts, but higher in 1970c by age 10/11 due to higher mortality in the 1958c.^56^ However, numbers with the outcome were small, so prevalence estimates were imprecise.

### MENTAL HEALTH

Mental health was the most compared outcome across cohorts. Most work focused on psychological distress (encompassing non-specific symptoms of anxiety and depression, and emotional discomfort) in adulthood, and emotional problems in adolescence. Mental health has been measured repeatedly throughout life in all cohort studies using questionnaires, partly helping to overcome problems related to changing diagnostic practices (including willingness to seek diagnosis) and stigma. However, there were differences in the questionnaires used across cohorts. Some studies have compared mental health “directly”, since some cohorts (e.g., 1958c and 1970c) have used the same questionnaires. Others have used harmonisation, calibration, or linking approaches to improve the cross-cohort comparability of mental health measures. In both cases, many studies have explicitly tested whether resulting measures were comparable by means of measurement invariance/equivalence testing.^23,57–60^

**Mental health in adulthood.** Studies comparing the 1946c, 1958c and 1970c (at specific ages and throughout adulthood) have consistently found that prevalence of psychological distress was higher in the 1970c compared to earlier born cohorts, using total questionnaire scores, factor scores, and binary ‘caseness’ measures of psychological distress based on clinically relevant cut-off scores.^20,23,25,35,39,57,58,60–64^ The 1970c had a higher prevalence of both low-level and high-level symptoms compared to the 1958c, and a lower prevalence of no symptoms.^65^ Comparisons with the 1946c were more challenging due to differences in the questionnaires used, requiring harmonisation.^66^ When all three cohorts were considered together, there was no strong evidence for a decline in mental health across 1946c and 1958c, but significantly higher levels of mental ill-health in 1970c relative to earlier cohorts.^58,60^

**Mental health in childhood and adolescence**. Comparisons of mental health in childhood and adolescence have occurred across the 1958c, 1970c, ALSPAC, and 2001c. Overall, cohort differences in mental health in childhood (age 7) were not apparent,^67^ but emerged when comparisons were made in adolescence (age 14-16). The 2001c consistently exhibited higher levels of mental ill-health (emotional problems, internalising symptoms, depressive symptomatology) in adolescence compared to earlier cohorts (1958c, 1970c, ALSPAC).^42,59,68,69^ Studies comparing internalising symptom levels between the 1958c and 1970c at age 16 using the parent-reported Rutter Scale generally found no evidence for a cohort difference.^23,59,61,70^ Several studies reported a higher prevalence of conduct disorders in 1970c compared to the 1958c.^23,59,61,70^ Evidence on externalising symptoms in later-born cohorts was more limited, though one study did find that mean levels of parent-reported conduct, hyperactivity, and peer problems were all higher in the 2001c compared to ALSPAC in adolescence.^68^

**Table 1.**
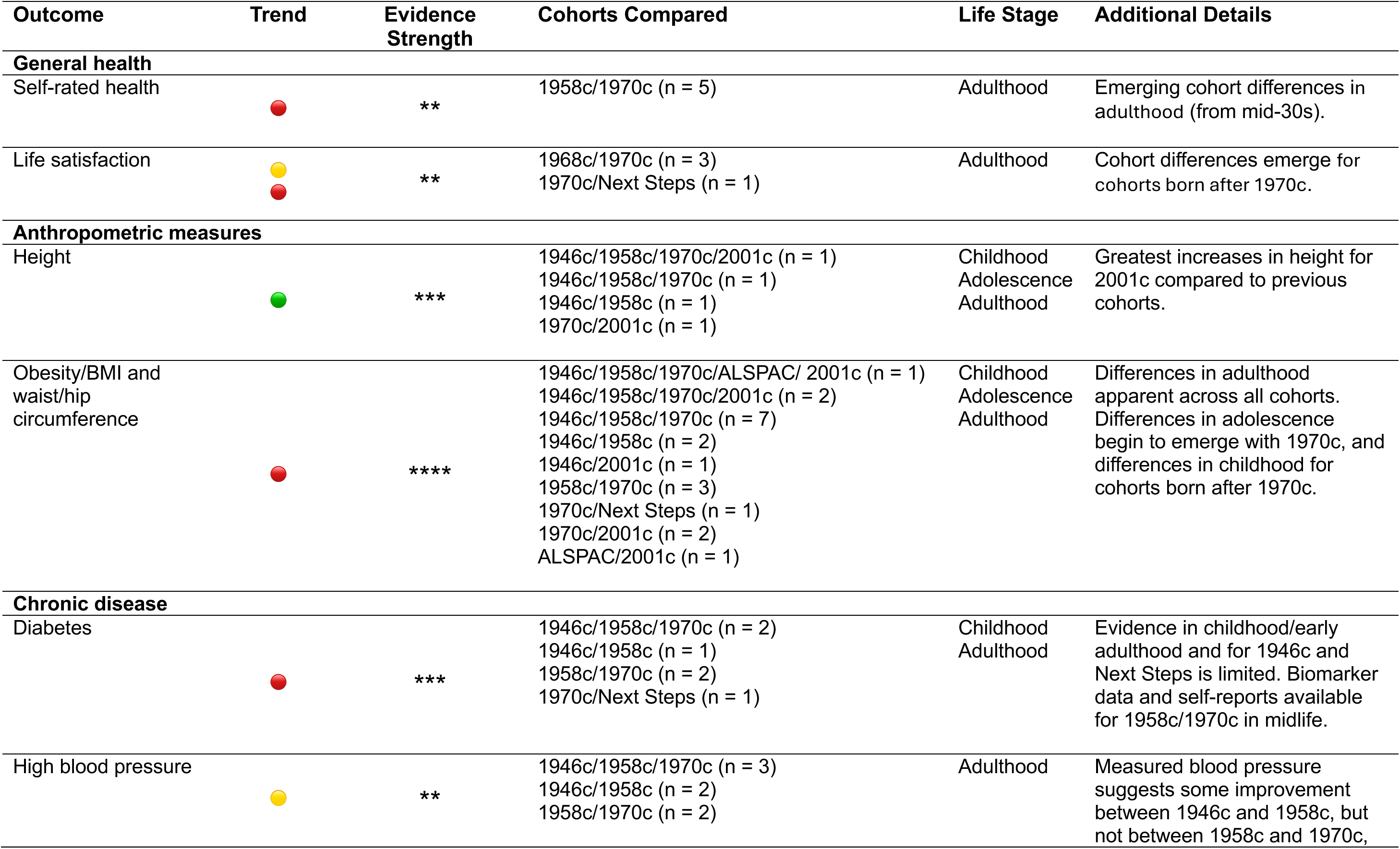
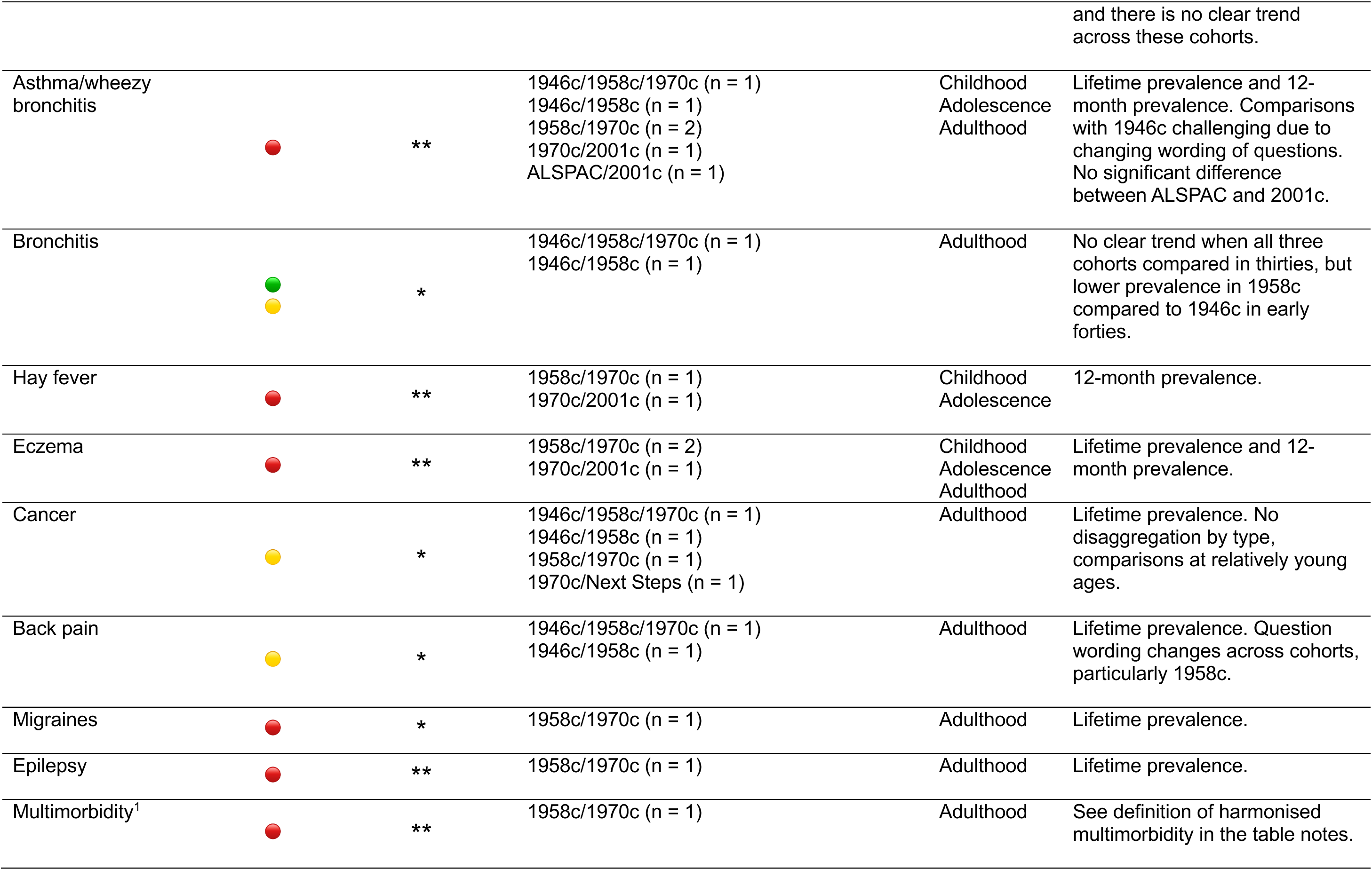
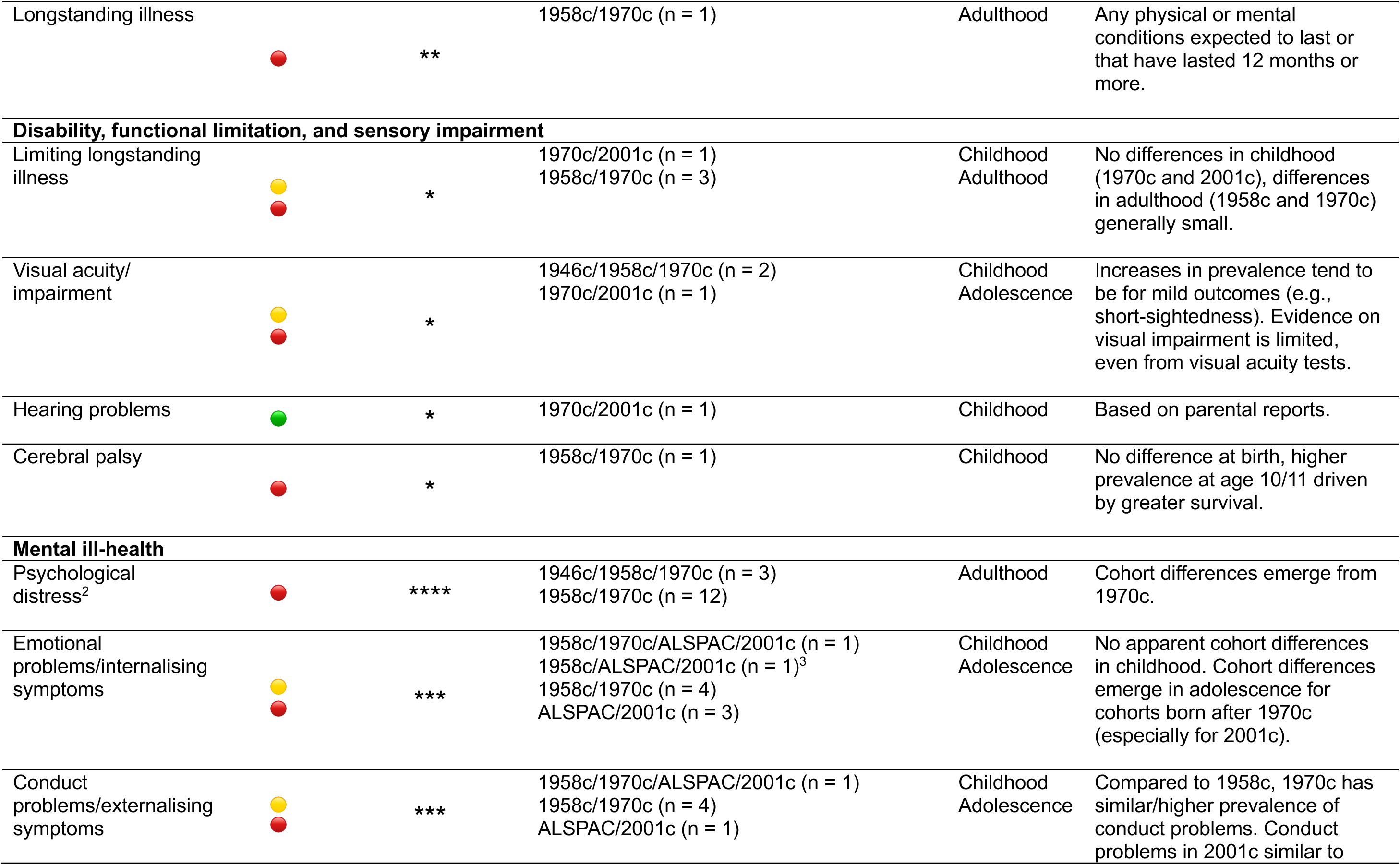
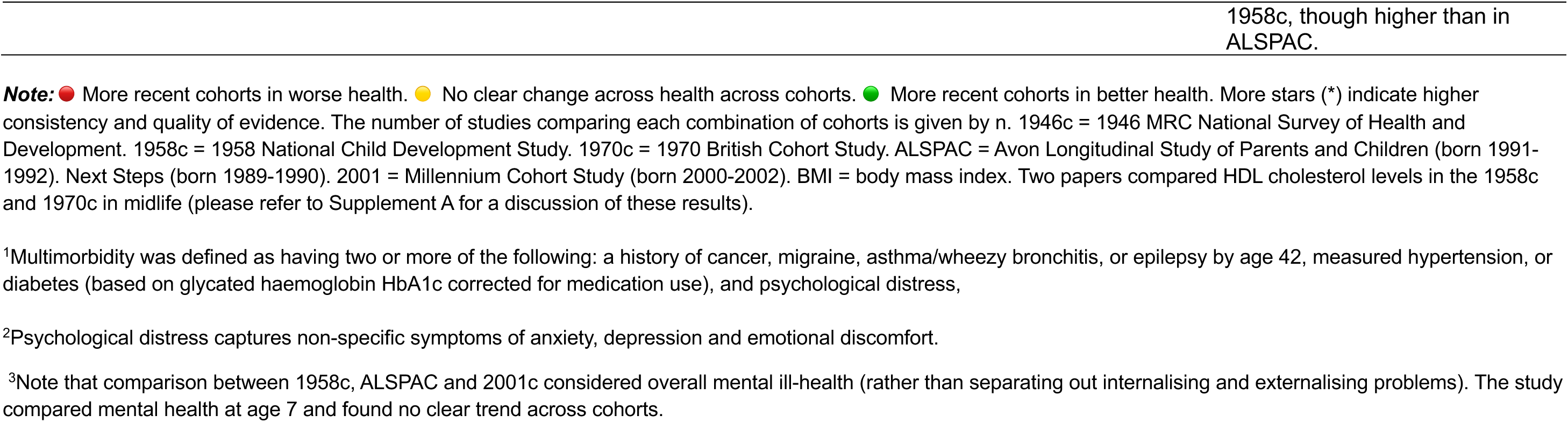
Summary of main findings.

## DISCUSSION

### SUMMARY OF THE EVIDENCE

Our findings suggest that the prevalence of physical and mental ill-health was similar or higher in more recently born cohorts for a range of self-reported and observer measured outcomes, when comparisons were made at the same age. Evidence was strong for earlier onset of obesity and mental ill-health, two outcomes that have been repeatedly measured since childhood and for which prevalence could be compared across the lifecourse. There was less evidence on other health outcomes, with most comparisons limited to specific ages, but overall, there was little evidence for declining prevalence of poor health across successive cohorts.

While this review only included publications comparing health across British birth cohort studies, findings from studies using different designs (cohort comparisons of age-adjusted prevalence, trends in age-specific and/or age-standardised prevalence) and data (repeated cross-sectional and longitudinal survey data, electronic health records, registry data) – with different assumptions and limitations – generally point in the same direction as our findings. A systematic review of population-based studies up to 2017 found that accounting for age, prevalence of most included outcomes increased or remained stable (including diabetes, asthma, and lower back pain) with the exception of dementia/Alzheimer’s disease.^71^ Two studies using pseudo-cohort designs similarly found no evidence for declines in the prevalence of most chronic disease in England for cohorts born since 1945.^72,73^ A summary of evidence from these studies can be found in Supplement A. Studies using actuarial methods have found that improvements in health expectancies in the United Kingdom have slowed or stalled since 2010, driven by worsening health in midlife.^4,74–76^ Since age-specific mortality and prevalence rates at younger ages factoring into period health expectancy calculations are contributed by successive cohorts, these findings appear to be consistent those of this review.

### INTERPRETATION OF THE FINDINGS

Two important questions emerge from our findings. The first is whether increasing prevalence of ill-health across cohorts is artefactual, that is, whether it reflects changes in how health is measured over time. The second is, if this trend is not entirely artefactual, whether it is driven by increasing survival or true worsening health. The relative importance of these explanations is likely to vary by health condition, and more research is needed to fully understand this.

#### Artefactual explanations for cohort trends

Many outcomes compared in this review were self-reported, and the generations represented by the British birth cohorts witnessed substantial improvements in the diagnosis, screening, treatment, and management of chronic disease. There have also been social changes (e.g., improvements in education) which have likely led to greater health literacy and awareness in more recent cohorts, affecting the extent to which medical diagnosis is sought out (e.g., changes in primary care attendance), how health is perceived (e.g., what is considered “normal” health), and willingness to report health problems (e.g., destigmatisation of mental ill-health).

Measuring change in total disease prevalence (including undiagnosed or unreported poor health) across cohorts is challenging, particularly for conditions which may be asymptomatic (e.g., hypertension, diabetes, early stages of cancer) and require screening to be diagnosed (and therefore known to the cohort member and self-reported). The introduction of screening programmes (e.g., National Health Service Health Check) and incentives in primary care (e.g., Quality and Outcomes Framework) means that members of more recent cohorts may be more likely to receive diagnoses than earlier cohorts at a given age. The extent to which different health conditions are self-reported once diagnosed also varies.^77,78^

Both self-reported and observed measures of diabetes and high blood pressure were available in the cohorts and evidence could be triangulated. The direction of trends was generally consistent with those from self-reports. Since biomarker data was not restricted to those presenting to the medical system, outcomes based on biomarkers capture both the diagnosed and undiagnosed (while self-reports capture only those aware of their condition). Evidence using other datasets and a pseudo-cohort approach have also found that the prevalence of observer-measured obesity, adiposity, and HbA1c across successive generations.^72,73,79,80^ For blood pressure and cholesterol, evidence is more mixed.

Comparing Generation X and Millennials, one study found that cholesterol and blood pressure levels were lower in the more recent cohort, with and without accounting for medication use.^80^ Another study comparing Baby Boomers to preceding generations found that, accounting for medication use, prevalence of high blood pressure, elevated HbA1c, and high total cholesterol had increased across cohorts.^72^ Biomarker evidence suggests that cohort trends in these conditions are not driven by changing diagnosis or reporting styles.

However, few conditions have such closely corresponding biomarkers, and nonresponse is typically higher for biomarker data. Self-reported outcomes therefore remain important and can inform planning for future healthcare demand.

A trend of worsening health is also observed for many outcomes where diagnosis is not necessary. For instance, criteria for defining obesity have remained the same over time. Since mental ill-health was measured using questionnaires on symptoms rather than by asking about diagnoses, prevalence estimates in the cohorts may more closely reflect the population prevalence of common mental health problems (including diagnosed and undiagnosed) and be less vulnerable to changing diagnostic practices or health-seeking behaviour across cohorts. Cohort differences in questionnaire item interpretation could still have led to artefactually higher prevalences of mental ill-health in more recent cohorts, but extensive work to harmonise measures of mental health and to explicitly test these measures for measurement invariance suggests this is unlikely to be the case.^23,57–60,81^ The same trend of worse health in the more recent cohort was also observed for self-rated health, another outcome that does not need to be diagnosed to be reported.

#### Improvements in survival and true increases in prevalence

Assuming cohort differences cannot be entirely explained by artefact, what mechanisms might explain this trend? Prevalence is a function of mortality and recovery (flows out of ill-health) and incidence (flows into ill-health). The impact of these mechanisms is likely to be different depending on the outcome considered.

Mortality rates have declined across successive cohorts. Those with poor health or at risk of poor health are more likely to survive to a given age in more recent generations. Increasing survival is likely an important driver of Generational Health Drift for conditions which have experiences rapid increases in survival (e.g., some cancer types). However, it is unlikely to explain results for other outcomes which are less strongly associated with mortality, especially at the relatively young ages when comparisons have occurred.

Several studies compared lifetime prevalence measures (capturing both those with the condition and who recovered) but comparisons using point-prevalence measures (excluding those who recovered) also tended to find that prevalence was higher in more recent cohorts, suggesting that these results are not entirely driven by higher recovery rates.

Increasing prevalence is also likely being partly driven by changes in age-specific incidence, with more recent cohorts experiencing earlier onset of ill-health on average (Figure 3). The birth cohort studies provide strong evidence of this for mental ill-health and obesity, with cohort differences emerging earlier in life across successive generations.

**Figure 3.**
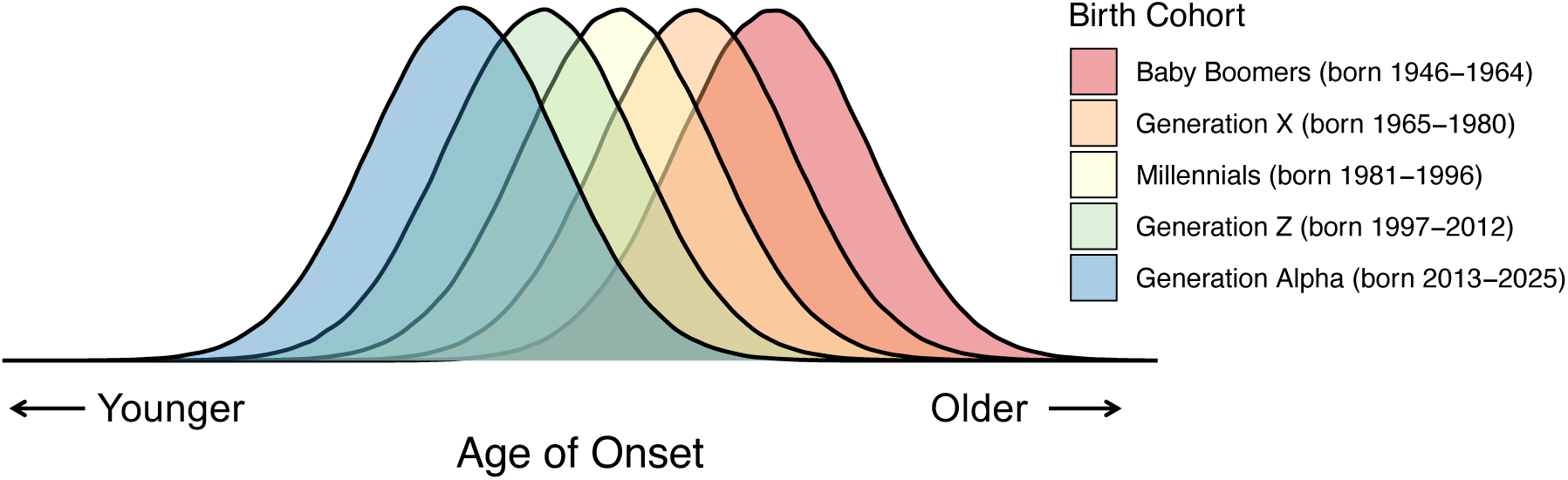
A graphical representation of declining age of onset of poor health across successive generations.

### STRENGTHS AND LIMITATIONS

The British birth cohorts were sampled to be representative of people born in Britain in specific years. In combination with their large size and long follow-up, this makes them unique tools for exploring cohort trends in health. Since individuals had to be born in Britain to be eligible for inclusion, the older cohorts are less ethnically diverse than the British population of the same age currently living (but not necessarily born) in Britain. However, recent evidence suggests that differences between these two populations may be minor for several important characteristics (e.g., mortality, longstanding illness, educational attainment).^82–85^ Strengths of the work synthesised in this review include the range of physical and mental health outcomes considered, triangulation across self-reported and observer-measured outcomes, and the repeated measurement of many outcomes across the lifecourse, offering opportunities to explore when and for which cohorts differences in prevalence emerged.

An important limitation of work using the British cohorts is that they suffer from attrition and nonresponse, like all longitudinal surveys. Many of the included studies restricted analytical samples to those who responded at similar ages, such that comparisons required additional assumptions to be made about the similarity of missing data mechanisms across cohorts.

More recent studies used inverse probability weighting and multiple imputation, which rely on the assumption that the probability of missingness depends only on observed data.

These techniques can successfully restore sample representativeness despite selective attrition when appropriate auxiliary variables are available, as has been shown in some of the included cohorts.^82,84,86,87^

It is also worth noting that while most studies are nationally representative, ALSPAC is a regional cohort study, and participants are less ethnically diverse and more socioeconomically advantaged than the national average.^14^ However, studies explicitly seeking to address this issue found no change in the direction of cohort differences once these differences were accounted for.^68^

Harmonisation of health variables is a major challenge in cross-cohort comparative research. Outcomes benefiting from harmonisation efforts (BMI, mental health) were the most frequently compared across cohorts.^88,89^ The extent to which retrospective harmonisation is possible varies by health condition, cohort and age. Shifting scientific priorities and the availability of better measurement instruments have changed questions on health asked in the cohorts. Further retrospective harmonisation efforts and continued prospective harmonisation of survey questionnaires will help to improve our understanding of cohort trends in health.

### FURTHER RESEARCH

Most studies compared prevalence of poor health at specific ages. Studies describing changing prevalence over age across cohorts were limited to a narrow set of outcomes (e.g., BMI, mental ill-health). Expanding the latter approach to other outcomes could provide insight into when (and for which cohorts) differences in health emerge.

As the British birth cohorts age, it will be possible to compare health across cohorts beyond midlife, into cohort members fifties, sixties, and seventies (when risk of ill-health increases sharply). Will cohort differences persist, narrow, or grow? What will be the implications of increasing obesity prevalence in these cohorts for more severe cardiovascular outcomes, disability, and cognitive decline – outcomes for which evidence is currently non-existent or limited?

More research is needed to understand the drivers of Generational Health Drift, which has occurred despite declines in smoking, increasing educational attainment, and improvements in early life material circumstances across post-war cohorts. The richness of the British birth cohort data offers unique opportunities to explore this question from a cross-generational lifecourse perspective (e.g., testing mediators of cohort-health associations, and how trends might differ for different groups). The cohorts could also contribute to better understanding the societal consequences of Generational Health Drift (e.g., implications for labour force participation and medical expenditure at older ages).

### IMPLICATIONS

Life expectancy has increased dramatically over recent decades.^90^ Provided mortality improvements are not reversed, a higher prevalence of ill-health in more recent generations at the same age suggests that more total years may be spent living with poor health.^91,92^ Since ill-health in this review was frequently operationalised as binary (yes/no), the possibility of dynamic equilibrium (more years in poor health, but fewer with severe health problems) cannot be ruled out,^93^ but our findings suggest that for included outcomes, morbidity compression (fewer years in poor health) will not occur across these cohorts.^94^

While several mechanisms could explain the Generational Health Drift, evidence from the British birth cohorts suggests that more recent cohorts are experiencing earlier onset of poor health for a several outcomes, particularly obesity and mental ill-health. “Drifting” backwards in health for younger generations implies that we are not reaching the biological limits of health improvement but seeing the consequences of preventable social and environmental exposures which have shaped population health over time and across generations. The Generational Health Drift has serious implications for policy, planning, and funding allocation to be able to support a greater number of people living with chronic health conditions.

## AUTHOR CONTRIBUTIONS

LG, DMA and GBP conceptualised the review, with contributions from AG and JBD. LG and DMA designed the search strategy and developed the study protocol, with contributions from GBP. LG searched the databases. LG, MD and YG screened, extracted, and quality assessed the data. LG wrote the first draft of the manuscript. All authors contributed to the interpretation of the findings and critically revised the manuscript.

## Data Availability

All studies used in this review have been published and are referenced. All data extracted from included studies is available in Supplement C. Information on studies excluded at the full-text screening stage is given in Supplement B.

