## Supplement A for "The Generational Health Drift: A Systematic Review of Evidence from the British Birth Cohort Studies"

### SUPPLEMENT A1: Cohort Studies Included

**Table S1.** Characteristics of the cohort studies included in the review.

| Cohort | Born | Region | Target population | Study design | Achieved initial sample | Sample size | Link to cohort profiles |
|---|---|---|---|---|---|---|---|
| NSHD | 1946 | England, Scotland, Wales | All births in one week of 1946. Subsequent follow-up of singleton births to married women. | No sampling at stage 1 (maternity survey). Then a stratified random sample of singleton births to married mothers from the maternity survey (all with fathers who had non-manual/agricultural jobs, 25% of those with fathers in manual jobs). | 82% of all eligible births included in maternity survey. | 5362 | Study website Cohort profile |
| NCDS | 1958 | England, Scotland, Wales | All live births in one week of 1958 | No sampling. Supplemented with immigrant children born in target week during childhood sweeps. | >98% of eligible births | ~17,500 (+ small number of immigrants added in childhood sweeps) | Study website Cohort profile |
| BCS70 | 1970 | England, Scotland, Wales | All live births in one week of 1970 | No sampling. Supplemented with immigrant children born in target week during childhood sweeps. | ~96% eligible births | ~16,500 (+ small number born in Northern Ireland and not followed up and small number immigrants added in childhood sweeps) | Study website Cohort profile Profile update |
| ALSPAC | 1991/92 | Avon (West of England, near Bristol) | All pregnancies for women resident in the area with an estimated delivery date between 1 April 1991 and 31 December 1992 inclusive. | Opportunistic sampling from women attending pre-natal clinics. Non-enrolled at phase 1 were re-invited to join when children were 7 and 8 years old. | 71.8% of eligible pregnancies recruited prenatally. 75.3% enrolled after subsequent recruitment rounds. | ~15,000 pregnancies (~14,800 live births) | Study website Cohort profile |
| Next Steps (formerly LSYPE) | 1989/90 | England | Pupils in Year 9 (aged 13/14) in English state and independent schools and pupil referral units in February 2004. | Two-stage probability proportional to size sampling with disproportionate stratification. Primary sampling units were maintained and independent schools and pupil referral units. From state schools, pupils selected from School Census with higher probability of selection for ethnic minority students. Otherwise, selected directly from the school roll. | 74% of issued sample | ~15,500 (+ ethnic minority boost at age 17 sweep) | Study website Cohort profile |
| MCS | 2000/02 | England, Scotland, Wales, Northern Ireland | All children who were born in the target period who alive and resident in UK at 9 months old, and on Child Benefit register (97% coverage by age 7 months; excludes asylum seekers and temporary residents). | Disproportionately stratified cluster sampling. In England, oversample electoral wards where >30% population Black/Asian in 1991 census, and in most deprived quartile of wards. In Wales, Scotland and Northern Ireland, oversample deprived electoral wards. Within strata, wards ordered by region and by size and sampled systematically. | ~72% of issued sample | ~19,000 (+ England-only boost at age 3 sweep). | Study website Cohort profile |

### SUPPLEMENT A2: Systematic Review Methods

A protocol for this review pre-registered with PROSPERO (CRD42024560326).[1] Any changes to the protocol are explicitly stated in the text below.

Four electronic databases (MEDLINE, EMBASE, Web of Science, PsycInfo, up to 25/06/2024) were searched for any pairwise combination of the six cohort studies, as well for terms indicating the inclusion of two or more British birth cohort studies in the abstract and title (Box S1). Search terms for health conditions were not specified to ensure health outcomes were not accidentally missed. Preprints were not excluded in an effort to comprehensively review the evidence, even if this had not been formally published. Only one paper eligible for inclusion was only available as a preprint (Righton *et al.*, 2024) at the time of data extraction. However, this paper was subsequently published in PLOS One in December 2024, and we updated the review to cite the published version of the paper.

**Box S1**. OVID search strategy used for EMBASE, MEDLINE and PsycInfo

1. (NSHD or "National Survey of Health and Development" or “1946 cohort” or “1946 British birth cohort” or “1946 British cohort” or “1946 birth cohort”)
2. (NCDS or “National Child Development Study” or “1958 cohort” or “1958 British birth cohort” or “1958 British cohort” or “1958 birth cohort”)
3. (BCS70 or “1970 British Cohort Study” or “1970 cohort” or “1970 birth cohort” or “1970 British birth cohort” or “1970 British Cohort” or “British Cohort Study” or BCS)
4. (ALSPAC or “Avon Longitudinal Study of Parents and Children”)
5. (“Next Steps” or “Longitudinal Study of Young People in England” or LSYPE)
6. (MCS or “Millennium Cohort Study”)
7. (two or three or four or five or “2” or “3” or “4” or “5”) and (“British birth cohorts” or “British cohorts” or “British birth cohort studies”)
8. (1 and 2) or (1 and 3) or (1 and 4) or (1 and 5) or (1 and 6) or (2 and 3) or (2 and 4) or (2 and 5) or (2 and 6) or (3 and 4) or (3 and 5) or (3 and 6) or (4 and 5) or (4 and 6) or (5 and 6) or 7

Titles and abstracts, and subsequently full texts, were screened by two reviewers (LG, and MD and YG acting as a pair) according to the criteria in Table S2 using Rayyan. Reviewers were blinded to one another’s decisions at both stages. Disagreements were resolved through discussion with a third reviewer.

During full-text screening, we noted that publications meeting the inclusion criteria in Table S2 fell into two categories: those aiming to explicitly compare health or associations between health and other variables across cohorts (Tier 1) and those that did not have this aim but provided a limited set of relevant descriptive statistics, usually when health was an exposure (Tier 2). Since the number of studies in this second group was large and evidence tended to be of lower quality, we additionally excluded Tier 2 publications if they conditioned the analytical sample on response to sweeps at different ages (e.g., respondents at age 42 in the 1970c, but at age 55 in 1958c), since the impact of attrition through mortality and loss to follow-up would affect the comparability of included cohort members. Note that this additional restriction was not stated in the protocol and was only applied at the full-text screening stage. As such, any Tier 2 publications excluded for this reason can be found listed in Supplement B (list of studies excluded during full-text screening with reasons for their exclusion).

**Table S2**. Study inclusion and exclusion criteria.

| | Inclusion | Exclusion |
|---|---|---|
| (1) | Published in any language. | |
| (2) | | Conference abstracts. |
| (3) | Included two or more of 1946c, 1958c, 1970c, ALSPAC, Next Steps or 2001c. | |
| (4) | Considered health as an exposure or an outcome. | Only considered health behaviours (smoking, drinking, diet, physical exercise), contacts with medical system (frequency of medical visits, hospital admissions), cognitive ability[1], or birthweight[2] as outcomes/exposures. |
| (5) | Compared cohorts at the same or similar ages (within 3 years)[3] or across a range of overlapping ages. | Pooled analyses where no cohort stratified results are presented. |
| (6) | Presentation of findings enabled prevalence to be compared across whole cohorts. Acceptable outcome measures included prevalence (estimated or observed), mean age of disease onset, and ratio measures (prevalence ratio, odds ratio, risk ratio, etc) from crude or minimally adjusted regression models (to avoid Table 2 fallacy) using cohort or birth year as the exposure. | Findings did not enable prevalence to be compared across cohorts. Analysis was restricted to specific subgroups whose size and/or composition has likely changed across cohorts (e.g., teenage mothers, married parents, university graduates) or only provided results stratified by subgroups whose size and composition has likely changed across cohorts. |

[1]Cognition was not included in the review. This was not explicitly stated in the study protocol. Given the ages at which cohort studies currently overlap (up to their mid-sixties), any available comparisons would have been on cognitive ability rather than age-related cognitive decline/dementia.

[2]Birthweight and gestational age were not included in the review. This was not explicitly states in the study protocol. Birthweight (and gestational age at birth) are known to be associated with higher risk of chronic disease in adulthood, but their relationship with health is non-linear (both low and high birthweight/gestational age can be risk factors), and cohort members are those who were born alive and survived the neonatal period. Comparisons of birthweight across the British birth cohorts are therefore more likely to reflect changing survival of low birthweight/preterm babies than true changes in the distribution gestational age and/or birthweight across all births.

[3]For some outcomes in childhood and adolescence, we used a narrower range. This was stated in the study protocol. For instance, period prevalence of asthma changes rapidly across childhood, so comparisons at age 5 and age 7 do not offer a strong age-specific comparison. These decisions were made based on full-text screening.

***Note:*** Papers could often be excluded for several reasons. We screened papers following the order given in Table S2 and retain the first reason for exclusion as the reason given in Figure 2 of the main manuscript.

Database searches yielded 1159 references, and an additional 4 references were identified through other means (searching reference lists, Google Scholar searches). After removing duplicates this equated to 494 publications. Titles and abstracts were screened (Cohen's Kappa = 0.9). A total of 105 publications were screened in full by two reviewers (Cohen's Kappa = 0.75). The final corpus consisted of 51 publications, of which 37 (73%) explicitly compared health or associations between health and other variables across cohorts (Tier 1), and 14 provided descriptive statistics on health outcomes at overlapping ages but did not explicitly aim to compare cohorts (Tier 2).

Data from included publications was extracted by LG and YG, with checks of a random 10% sample performed by MD and YG to ensure reliability. Fields extracted included key bibliographic information, details on the health outcomes compared and their measurement in each cohort, and information of the choice of analytical sample and how missingness and attrition were addressed.

Risk of bias of included studies was assessed by one reviewer (LG) and a random 10% sample was checked by MD and YG. This is a slight change from the protocol, where it was suggested that two researchers would independently carry out the quality assessment. Studies were assessed using the Joanna Briggs Institute Prevalence Studies Criterion.[2] A version of the assessment tool with additional information on how it was applied to these studies is shown in Table S3 below. Risk of bias was assessed at the individual publication level (and reported in Supplement C) and discussed at the level of the outcome.

Owing to the range of health outcomes assessed, and cohorts and ages assessed, results were findings narratively, with emphasis given to the direction of the trend across cohorts and the consistency of findings across studies. In the study protocol, we suggested that results would be summarised separately for observer-measured and self-reported outcomes. However, since there was usually close correspondence between observer-measured outcomes and a particular self-reported condition (e.g., measured BMI and obesity, HbA1c and diabetes, measured and self-reported high blood pressure), we instead described our results by health condition. A limitation of this approach was that, while for some conditions there were multiple studies that provided evidence for the same cohort/age combinations, allowing us to assess the consistency of findings across studies, this was not the case for most outcomes. However, for the same reasons (variety of outcomes and cohort/age combinations), the review arguably paints a comprehensive picture of changes in population health across cohorts.

**Table S3.** Risk of bias assessment criteria

| | | Yes | No | Unclear | N/A |
|---|---|---|---|---|---|
| 1 | Was the sample representative of the target population?<br>*In these studies, the target population is usually defined as those alive and living in Britain/the United Kingdom at the time of data collection. We paid specific attention to whether analytical samples conditioned on the same target population (i.e., survival to the same age), and whether authors used any techniques to ensure that the analytical sample was representative of the target population (e.g., multiple imputation back to the target population, use of inverse probability weights for non-response).* | | | | |
| 2 | Were study participants recruited in an appropriate way?<br>*This criterion was less relevant for quality assessment of individual publications, since all included publication used data from the same collection of longitudinal datasets. Information on study recruitment and baseline response rates are provided in Table 1 in the main manuscript.* | | | | |
| 3 | Was the sample size adequate?<br>*We considered how common health outcomes were and whether the size of the cohort and of the analytical sample was sufficient to adequately capture prevalence.* | | | | |
| 4 | Were the study subjects and the setting described in detail?<br>*Since the publications included in this review used the same set of longitudinal studies, we focused more on whether authors adequately described any additional restrictions made to the study cohort (e.g., only respondents to data collection sweep X) and whether they discussed how these decisions might impact prevalence estimates.* | | | | |
| 5 | Was the data analysis conducted with sufficient coverage of the identified sample?<br>*Were analyses restricted to those with complete data? If so, what proportion of the target population was included in analyses? Was a principled approach to missing data (e.g., multiple imputation) used?* | | | | |
| 6 | Were objective, standard criteria used for the measurement of the condition?<br>*Was the outcome measured by an observer (e.g., biomarker data, weight and height measured during medical examination), or was the outcome self-reported by the cohort member (or their parent in childhood data collection sweeps)? For self-reported outcomes, were standard, validated questionnaires used (e.g., Malaise Inventory)? For self-reported outcomes, were survey questions phrased similarly in both studies?* | | | | |
| 7 | Was the condition measured reliably?<br>*For observer-measured outcomes, was a standard protocol followed to collect data, and to what extent were these protocols comparable across studies? For self-reported outcomes, was any additional work done to establish similarity of reporting styles across cohorts (e.g., establish measurement invariance) or examine/discuss possible impact of mode effects?* | | | | |
| 8 | Was there appropriate statistical analysis?<br>*In cases where studies with complex sampling designs were used (i.e., 1946c, Next Steps, 2001c), did analyses appropriately account for these sampling designs (clustering, stratification)?* | | | | |
| 9 | Are all important confounding factors/subgroups/differences identified and accounted for?<br>*This review focused on measures of prevalence. An acceptable outcome measure was relative measures (risk ratio, rate ratio, odds ratio) provided cohort or year of birth was the exposure, health was the outcome, and regressions were unadjusted or minimally adjusted to avoid Table 2 fallacy.* | | | | |
| 10 | Were subpopulations identified using objective criteria?<br>*We did not consider this point in the quality assessment, since papers which presented results only for subgroups or stratified by subgroup whose composition was susceptible to be different across cohorts (e.g., educational status, marital status) were excluded during the screening stage. As such, the only subgroup analyses in publications included in the review were by sex at birth.* | | | | |

**Note:** We assessed the quality of included studies using criteria based on the Joanna Briggs Institute Prevalence Critical Appraisal Tool (Munn, Moola, Riitano & Lisy, 2014, *Int J Health Policy Manag,* 3(3): 123-128, doi: 10.15171/ijhpm.2014.71). In italics, we draw attention to specific aspects of the papers that we considered, tailored to the research question of the review (i.e., comparability between studies, longitudinal nature of the cohorts).

#### SUPPLEMENT A3: HDL Cholesterol

Two studies have compared levels of high-density lipoprotein (HDL) cholesterol in the 1958c and 1970c in midlife (ages 44-48).[3,4] Accounting for changes in medication use and applying sex-specific cut-offs, both studies found that the prevalence of low HDL cholesterol was significantly higher in the 1970c. However, the studies provided substantially different cohort-specific estimated prevalences, possibly because of a large amount of missing data on biomarkers and differences in how the analytical cohorts were defined.

HDL cholesterol is a regular feature of blood lipid profiles and there are guidelines for what are considered “healthy levels” of HDL cholesterol in the United Kingdom (>1 mmol/L for men and >1.2 mmol/L for women).[5] Low HDL cholesterol is associated with worse cardiovascular outcomes and with higher mortality in large, longitudinal observational studies.[6] More recent studies have challenged the idea, however, that the association between HDL cholesterol and cardiovascular outcomes is linear (i.e., that higher HDL cholesterol is universally protective against cardiovascular disease), suggesting instead that the relationship may be U-shaped, with both low and high levels of HDL cholesterol being associated with worse outcomes.[6,7] Our review is primarily concerned with comparing the prevalence of markers of poor health across cohorts. Given that low HDL cholesterol can serve as a predictive biomarker for poor health outcomes like cardiovascular disease, the higher prevalence of low HDL in the 1970c could be interpreted as another finding consistent with “Generational Health Drift”.

It is important to note, however, that there is an ongoing debate over the causal role of HDL cholesterol in cardiovascular disease. The biological mechanisms linking HDL cholesterol levels and cardiovascular outcomes are not yet fully understood.[8] Interventions to increase HDL levels do not appear to improve cardiovascular outcomes,[6] and Mendelian randomisation studies have raised questions over the role of unmeasured confounding in the negative association between HDL cholesterol and poor health outcomes seen in observational studies.[8]

The relationship between HDL cholesterol and cardiovascular disease is therefore not straightforward, and it is for this reason that we discuss these findings in depth here rather than in the main manuscript.

### SUPPLEMENT A4: Evidence From Other Study Designs

We conducted a rapid review of recent evidence from three study designs which yield evidence on how prevalence has changed accounting for age.

- **Pseudo-cohort approaches** use data from repeated cross-sectional studies or multiple waves of longitudinal studies to construct exposure groups (of different subjects or the same subjects observed several times) based on birth year. Within each exposure group, participants are observed at different ages, and the prevalence of the outcome is compared by statistically adjusting for age.
- **Period trends in age-specific prevalence** can also indirectly yield evidence on how prevalence has changed across cohorts, provided trends are observed across a large enough timespan and age-groups are narrow enough for sufficiently different cohorts to pass through the 'age-specific' window.
- **Period trends in age-standardised prevalence** can similarly produce evidence on how health has changed across cohorts by applying age-specific rates to a reference age-distribution, provided, as above, that the period of observation is long enough, and age-groups are narrow enough for different cohorts to be captured within each 'age-specific' window. One challenge of this design is that cohort trends in prevalence could be obscured there has been a change in the direction of prevalence trends (e.g., previously declining across cohorts born up to 1945, followed by increases across cohorts born since then).

Below, we summarise the main findings from this rapid review, which updated findings from a previous systematic review by Gondek and colleagues.[9] This review synthesised evidence published up to 2017 on trends in health expectancy and prevalence of multiple health outcomes in Britain since 1946, with the overall aim of interpreting these findings through an expansion/compression of morbidity framework. We replicated the search strategy used in this review, focusing on prevalence, and searched for publications since 2017. We summarise the evidence here, weaving together findings from the review by Gondek and colleagues with those from our updated literature search.

The prevalence of poor **self-rated health** (on a scale from "poor" to "excellent") increased between the 1980s and 2000s based on a previous systematic review.[9] Using pseudo-cohort approach and adjusting for age, a study using Health Survey for England data found that the odds of poor self-rated health increased for both men and women across successive cohorts of 25 to 64-year-olds born between 1945 and 1980.[10] However, another study using General Household Survey data found that, for pseudo-cohorts of adults aged 30-59 born 1920 to 1970, the age-adjusted probability of having "not good" self-rated health remained stable across cohorts for both men and women.[11] In these same studies, the prevalence of longstanding illness remained stable for women, and there was some evidence for decreasing prevalence among men.

A systematic review on trends in the age-specific and age-standardised prevalence of major chronic non-communicable diseases and disability in the UK up to 2017 concluded that the prevalence of **lung cancer, diabetes, chronic obstructive pulmonary disease, asthma, cirrhosis, migraine, and backpain** has increased since 1946.[9] Prevalence of **coronary heart disease** and **stroke** were found to have remained stable, and prevalence of **dementia/Alzheimer's disease** was the only condition included in the review that showed consistent evidence for a decline. We replicated the search strategy used in this review paper to find publications describing trends in prevalence of disease and disability since 2017.

Since 2017, research on trends in age-specific and age-standardised disease prevalence has primarily leveraged large administrative datasets from primary and secondary healthcare. However, findings generally continue to support the conclusions of the systematic review. Studies using electronic health records data have found that the age-standardised prevalence of **heart failure** in adults aged ≥16 years remained stable between 2002 and 2014,[12] consistent with stability in coronary heart disease prevalence noted in the review, and the age-standardised prevalence of **type 2 diabetes** increased between 2004 and 2016.[13] Using the same data, the age-standardised prevalence of **osteoarthritis** in adults aged ≥20 years increased between 1997 and 2017.[14] The age-standardised prevalence of **dementia** fell across two cohorts of adults aged ≥65 years surveyed in 1991 and 2011.[15]

Studies using more 'direct' approaches than comparisons of age-standardised prevalence, such as pseudo-cohort approaches or trends in age-specific prevalence, also support the idea that more recently born cohorts, particularly those born since World War II, are experiencing worse health at the same age. Adjusting for age, prevalence of **self-reported diabetes, cardiovascular disease,** and **high blood pressure** (in men), **overweight** BMI, and **high HbA1c** increased across pseudo-cohorts of English adults aged 25-64 born 1945-1980.[10] Another study compared the prevalence of cardiometabolic risk factors among Generation X (born 1965-1980) and Millennials (born 1981-1996) in England at ages 20-34 years and found that the prevalence of **obesity** (in women) and high HbA1c increased across generations, while the prevalence of high blood pressure and **high cholesterol** declined.[16] Comparing cohorts of Scottish adults born in the 1930s, 1950s, and 1970s, **adiposity** (based on BMI and waist-to-height ratio) increased fastest across the lifecourse in the most recently born cohort.[17] A recent pseudo-cohort study using data from the English Longitudinal Study of Ageing (ELSA) found that compared to those born in 1936-1945, members of later born cohorts had a higher prevalence of ever having had **heart problems, cancer, lung problems, self-reported high cholesterol, self-reported diabetes and self-reported high blood pressure**, adjusting for age and gender.[18] There was also some suggestion that previous declines in **disability** had stalled, and that **grip strength** had declined (additionally adjusting for height and weight). Trends in biomarkers (**HbA1c, total cholesterol** and **systolic blood pressure** and **diastolic blood pressure**) accounting for medication corroborated trends found for self-reported diabetes, high blood pressure, and high cholesterol, and **obesity** prevalence also increased across cohorts.[18]

Trends in the prevalence of **limiting longstanding illness** (LLI) and disability in studies up to 2017 were mixed.[9] While nearly all studies found that the number of years spent with LLI or disability increased across cohorts, some studies found this increase to be driven by more rapid increases in life expectancy than age of disability onset, while other studies found that prevalence of disability or LLI was higher in more recent cohorts. A similar conclusion can be drawn from studies published since 2017. The prevalence of LLI declined across pseudo-cohorts of adults aged 25-64 years born 1945-1980.[10] Age-standardised prevalence of **disability** in adults aged ≥65 years in England declined between 2004 and 2014, while prevalence of severe disability at aged ≥80 years remained stable.[19]

Evidence from sources other than the British birth cohort studies also support the idea of a generational drift in **mental health.** A systematic review concluded that there was evidence of an increasing trend across generations in the prevalence of affective disorders in children and adolescents in the UK.[20] Two studies using data from the Adult Psychiatry Morbidity Survey (APMS), a repeated cross-sectional study, found no overall trend of worsening prevalence of common mental health disorders or depression for cohorts born between 1939 and 1981 in England.[21,22] However, in both instances, there was evidence for a step-increase in the prevalence of poor mental health and depression outcomes among those born in the mid-1940s and early 1950s onwards. Other studies have not found evidence of deteriorating mental health across generations. A study comparing parent- and teacher-reported mental health outcomes in 11-year-old children assessed in 1999, 2004, and 2012 did not find consistent worsening mental health across these generations.[23] Another study found that age-specific ‘caseness’ (a strong likelihood of experiencing a common mental health disorder) had not significantly changed for men born in 1916-1930, 1931-1945, 1946-1960, and 1961-1975, whereas it had improved for women.[24]

### SUPPLEMENT A5: PRISMA Reporting Guidelines

| Section and Topic | Item # | Checklist item | Location where item is reported |
|---|---|---|---|
| **TITLE** | | | |
| Title | 1 | Identify the report as a systematic review. | 1 |
| **ABSTRACT** | | | |
| Abstract | 2 | See the PRISMA 2020 for Abstracts checklist. | 150-word unstructured abstract<br>(some additional information provided in Research in Context box) |
| **INTRODUCTION** | | | |
| Rationale | 3 | Describe the rationale for the review in the context of existing knowledge. | 2 |
| Objectives | 4 | Provide an explicit statement of the objective(s) or question(s) the review addresses. | 3 |
| **METHODS** | | | |
| Eligibility criteria | 5 | Specify the inclusion and exclusion criteria for the review and how studies were grouped for the syntheses. | Suppl A2: Table S2 |
| Information sources | 6 | Specify all databases, registers, websites, organisations, reference lists and other sources searched or consulted to identify studies. Specify the date when each source was last searched or consulted. | 3 |
| Search strategy | 7 | Present the full search strategies for all databases, registers and websites, including any filters and limits used. | Suppl A2, Box S1 |
| Selection process | 8 | Specify the methods used to decide whether a study met the inclusion criteria of the review, including how many reviewers screened each record and each report retrieved, whether they worked independently, and if applicable, details of automation tools used in the process. | Suppl A2 |
| Data collection process | 9 | Specify the methods used to collect data from reports, including how many reviewers collected data from each report, whether they worked independently, any processes for obtaining or confirming data from study investigators, and if applicable, details of automation tools used in the process. | Suppl A2 |
| Data items | 10a | List and define all outcomes for which data were sought. Specify whether all results that were compatible with each outcome domain in each study were sought (e.g. for all measures, time points, analyses), and if not, the methods used to decide which results to collect. | Protocol, Table S2 |
| | 10b | List and define all other variables for which data were sought (e.g. participant and intervention characteristics, funding sources). Describe any assumptions made about any missing or unclear information. | Protocol, Suppl A2 |
| Study risk of bias assessment | 11 | Specify the methods used to assess risk of bias in the included studies, including details of the tool(s) used, how many reviewers assessed each study and whether they worked independently, and if applicable, details of automation tools used in the process. | 3-4, Suppl A2 |
| Effect measures | 12 | Specify for each outcome the effect measure(s) (e.g. risk ratio, mean difference) used in the synthesis or presentation of results. | Suppl A2: |

| Section and Topic | Item # | Checklist item | Location where item is reported |
|---|---|---|---|
| | | | Table S2 |
| Synthesis methods | 13a | Describe the processes used to decide which studies were eligible for each synthesis (e.g. tabulating the study intervention characteristics and comparing against the planned groups for each synthesis (item #5)). | Suppl A2 |
| | 13b | Describe any methods required to prepare the data for presentation or synthesis, such as handling of missing summary statistics, or data conversions. | N/A |
| | 13c | Describe any methods used to tabulate or visually display results of individual studies and syntheses. | N/A |
| | 13d | Describe any methods used to synthesize results and provide a rationale for the choice(s). If meta-analysis was performed, describe the model(s), method(s) to identify the presence and extent of statistical heterogeneity, and software package(s) used. | 4 |
| | 13e | Describe any methods used to explore possible causes of heterogeneity among study results (e.g. subgroup analysis, meta-regression). | N/A |
| | 13f | Describe any sensitivity analyses conducted to assess robustness of the synthesized results. | N/A |
| Reporting bias assessment | 14 | Describe any methods used to assess risk of bias due to missing results in a synthesis (arising from reporting biases). | Suppl A2 |
| Certainty assessment | 15 | Describe any methods used to assess certainty (or confidence) in the body of evidence for an outcome. . | Suppl A2 |
| **RESULTS** | | | |
| Study selection | 16a | Describe the results of the search and selection process, from the number of records identified in the search to the number of studies included in the review, ideally using a flow diagram. | 4 |
| | 16b | Cite studies that might appear to meet the inclusion criteria, but which were excluded, and explain why they were excluded. | Suppl B |
| Study characteristics | 17 | Cite each included study and present its characteristics. | Suppl C |
| Risk of bias in studies | 18 | Present assessments of risk of bias for each included study. | Suppl A2 |
| Results of individual studies | 19 | For all outcomes, present, for each study: (a) summary statistics for each group (where appropriate) and (b) an effect estimate and its precision (e.g. confidence/credible interval), ideally using structured tables or plots. | Suppl C |
| Results of syntheses | 20a | For each synthesis, briefly summarise the characteristics and risk of bias among contributing studies. | 5-9, Suppl C |
| | 20b | Present results of all statistical syntheses conducted. If meta-analysis was done, present for each the summary estimate and its precision (e.g. confidence/credible interval) and measures of statistical heterogeneity. If comparing groups, describe the direction of the effect. | N/A |
| | 20c | Present results of all investigations of possible causes of heterogeneity among study results. | N/A |
| | 20d | Present results of all sensitivity analyses conducted to assess the robustness of the synthesized results. | N/A |
| Reporting biases | 21 | Present assessments of risk of bias due to missing results (arising from reporting biases) for each synthesis assessed. | Suppl A2 |
| Certainty of evidence | 22 | Present assessments of certainty (or confidence) in the body of evidence for each outcome assessed. | 5-9, Suppl C |

| Section and Topic | Item # | Checklist item | Location where item is reported |
|---|---|---|---|
| **DISCUSSION** | | | |
| Discussion | 23a | Provide a general interpretation of the results in the context of other evidence. | 14 |
| | 23b | Discuss any limitations of the evidence included in the review. | 16 |
| | 23c | Discuss any limitations of the review processes used. *A limitation of this review is that results had to be summarised narratively, due to the wide range of outcomes, cohorts and ages considered. While for some conditions there were multiple studies that provided evidence for the same cohort/age combinations, allowing us to assess the consistency of these findings, this was not the case for most outcomes.* | Suppl A2 |
| | 23d | Discuss implications of the results for practice, policy, and future research. | 17 |
| **OTHER INFORMATION** | | | |
| Registration and protocol | 24a | Provide registration information for the review, including register name and registration number, or state that the review was not registered. | 3 |
| | 24b | Indicate where the review protocol can be accessed, or state that a protocol was not prepared. | 3 |
| | 24c | Describe and explain any amendments to information provided at registration or in the protocol. | Suppl A2 |
| Support | 25 | Describe sources of financial or non-financial support for the review, and the role of the funders or sponsors in the review. | 18 |
| Competing interests | 26 | Declare any competing interests of review authors. | 18 |
| Availability of data, code and other materials | 27 | Report which of the following are publicly available and where they can be found: template data collection forms; data extracted from included studies; data used for all analyses; analytic code; any other materials used in the review. | Suppl A2, Suppl B, Suppl C |

*From:* Page MJ, McKenzie JE, Bossuyt PM, Boutron I, Hoffmann TC, Mulrow CD, et al. The PRISMA 2020 statement: an updated guideline for reporting systematic reviews. BMJ 2021;372:n71. doi: 10.1136/bmj.n71.
This work is licensed under CC BY 4.0. To view a copy of this license, visit https://creativecommons.org/licenses/by/4.0/
